# Origin of HIV/AIDS and Vaccine Failure: Simulation of the Role of the Adjuvant Activation Hypothesis

**DOI:** 10.1101/2021.01.13.21249768

**Authors:** H.C. Grassi, E.D.J. Andrades, V.J. Andrades-Grassi

## Abstract

Human Immunodeficiency Virus (HIV) is the etiological agent for Acquired Immunodeficiency Syndrome (AIDS). It is possible that vaccine failure could be related to the events involved at the origin of HIV/AIDS. In this work the role of the adjuvant activation hypothesis on the origin and on the failure of vaccines, as well as other effects is evaluated by means of a simulation using a mathematical analysis, differential equations and an Excel spreadsheet. The results show that the adjuvant activation alters the viral load and the cellular and humoral Immune Response. Under certain conditions it was possible to show how the adjuvant activation could have promoted the origin of the HIV/AIDS pandemic and also, as a consequence of the SIV adaptation to human beings at the origin, the failure of present day vaccine trials. Other effects such as Immunotolerance and Antibody Dependent Enhancement (ADE) were shown. This study provides a means to examine other effectors in order to suggest therapeutic alternatives. In this case passive immunization in combination with anti-retroviral therapy showed an acceptable adaptation to the conditions tested. It is concluded that the methodological strategy of this work may be useful for the analysis of the adjuvant activation hypothesis as well as other effects, interactions and new proposals, such as thermodynamics of HIV infections.

## Introduction

The events that led to the origin of HIV infections and the origin of the subsequent AIDS pandemic, are probably conditioning the future events such as vaccine failure, disease progression in infected individuals and the dual interaction of the virus with CD4+ T lymphocytes (Grassi & Andrade, 2001). This refers to the dual compromise of activated CD4+ T lymphocytes first as helper cells with an important role in the Cellular and Humoral Immune Response and second as the main targets as host cells for HIV reproduction and reservoir (Finzi & Siliciano, 1998). Then, progression of the infection to the AIDS syndrome is also the product of another dual compromise of the activated helper lymphocytes. After infection and integration, CD4+ T cells can either take the pathway of the memory state remaining as a reservoir for the virus or the pathway for virus production (Zhou, Zhang, Siliciano, & Siliciano, 2005).

Evolutionary Darwinian points of view have been considered for the origin of HIV/AIDS (Talenti, 2005) (Sharp & Hahn, The Evolution of HIV-1 and the Origin of AIDS, 2010) (Sharp & Hahn, Origins of HIV and the AIDS Pandemic, 2011). However, confusion over the origin of the virus and the origin of the epidemics have been set (Marx, Apetrei, & Drucker, 2004). In the Adjuvant Activation Hypothesis (Grassi & Andrade, 2001) we propose that massive, extensive and intensive vaccination in the exposed population of Equatorial Africa played a crucial role in the selection of a sufficiently aggressive strain of HIV that progressed to AIDS. In spite of the fact that the authors consider the very important role that vaccines have played in maintaining human health and in prevention of infectious diseases, it is important to also consider collateral effects that may have occurred on the human population. Our proposal (Grassi & Andrade, 2001) refers to the Adjuvant Activation Effect (AAE) on the adaptation of the zoonotic virus (from SIV to HIV) to humans and the rise of the pandemic in relation to the activation of the Immune System of more than 30 million people due to anthropogenic activities of massive, extensive and intensive vaccination in Equatorial Africa, from the 1940’s to the 1970’s. This hypothesis anticipates that in evolutionary terms, viral particles that represent variant viruses would have been positively selected because some of the variants would infect newly activated T-cell clones as the massive, extensive and intensive vaccination took place. This would imply that there would have been an evolutionary natural positive selective pressure on strains of HIV, in zoonotic transition with a high mutation rate which were adapting to humans, and would have provided a higher number of multiple variant viruses and consequently a higher number of new activatable clones of CD4+ T cells. This AAE may have promoted the adaptation of the simian virus to the human Immune System. This hypothesis predicts that if this activation was the driving force for the adaptation of the simian virus to humans, then any activation of immune cells particularly CD4+ T cells could favor virus infection, hence the difficulty in designing vaccines using the whole virus or its parts. Furthermore, not only is HIV responsible for the activation of the CD4+ T lymphocytes, but other infections, conditions and vaccine challenges, as well as variants of HIV may contribute as stimulators of the Immune System in what has been referred to as AAE. This adjuvant activation hypothesis, predicted in 2001 that traditional vaccines for HIV/AIDS would fail and it anticipated the effect of secondary activations on the progression of the infection to the AIDS syndrome. The afore mentioned dual compromises of the HIV-CD4+ T cell interaction are also important in the development of vaccines which has been guided by the traditional search of promoting an anamnestic response which necessarily results in a rapid activation of CD4+ T lymphocytes among other components of the Immune System. Again, these cells can be either target for virus integration and replication or on the other hand, helper cells for the elimination of the virus or of infected cells (Finzi & Siliciano, 1998). Fauci et al (Fauci, Marovich, Dieffenbach, Hunter, & Buchbinder, 2014) consider that the level of protection seen with a vaccine can be viewed as the balance between the responses to the vaccine that lead to susceptibility to infection and by the responses that favor protection, although it is clear that vaccination may enhance immune activation, particularly of homologous CD4+ T cells (Fauci, Multifactorial Nature of Human Immunodeficiency Virus Disease: Implication for Therapy, 1993).

This paper aims to show the dual functions of CD4+ T cells as helper/virus host cells and on the other hand, the activated cells as HIV productive/reservoir and their implication in the AAE, using a dynamic Excel based model with 9 differential equations. Therefore, the main focus is set on activated CD4+ T cells and their interaction with other cells and with HIV. The original working hypothesis (Grassi & Andrade, 2001) is that this AAE drove in part the SIV to HIV transition, it was implicated in the origin of the pandemic and it is involved in AIDS progression and vaccine failure. In this work the authors do not intend to go against the use of vaccines nor to describe exhaustively the Immune System. Instead some of its selected elements are presented for the purpose of simulating the AAE in individuals which have a cellular population dynamic which is probably different from that of the population in which SIV evolved to HIV in the process of adapting to humans.

## METHODS

Interaction of the Nodes with HIV and the AAE

Table 1 has the description of the Nodes involved in this study. It should be noted that the main focus is on activated CD4+ T cells (Node N), either homologous or heterologous to HIV. Upon encounter with HIV, a fraction of CD4+ T cells will become activated (in a response as helper T cells) and a fraction of them could be infected with incorporation of the retro transcribed viral DNA into the host genome (Node E). Non-infected CD4+ T cells may subsequently enter the memory state (Node D) that later may also become infected and converted to Node M. Both, infected memory and activated infected (Node E) contribute to the productive Node M which represents the main subset of cells that produce the virus (see also Figure 1). This is the main focus of our approach because it contains the paired functions of CD4+ T cells as “activated-helper”/” activated-infected” and “activated-virus production”/”memory-virus latency”. Figure 1 shows the interaction of the cells that are relevant for the AAE. The upper nodes represent these 4 different states of CD4+ T cells, activated non-infected (Node N), memory non-infected (Node D), activated infected (Node E) and memory infected and productive (Node M). These nodes are affected (activated) by the presence of the virus (central node, V) and by CD8+ T and other cytotoxic cells which will contribute in the elimination of virus infected cells. Two states of these cells, naive (Node T) and activated (Node L) are considered. In addition, the extracellular virus will be controlled by the antibody titer, thus B lymphocytes in naive (Node C) and activated plasma cells (Node W) are considered. Finally, since the role in the AAE for CD4+ T cell activation is played both, by the HIV virus itself as well as by other infections and physiological or pathological conditions, an effector (A) for the incorporation of different AAE stimuli that activate these cells is included together with the HIV virus node. It should be noted that at the onset of the HIV infection, the activated CD4+ T cells will correspond to the specific homologous HIV subset, however as the infection progresses, different subsets of heterologous CD4+ T cells, will be incorporated, according to the AAE hypothesis (Grassi & Andrade, 2001). These stimuli include HIV variant viruses as well as microorganisms and other physiological and pathological conditions. Other effectors that may play a role on the viral count are also included as (B).

**Table 1.**
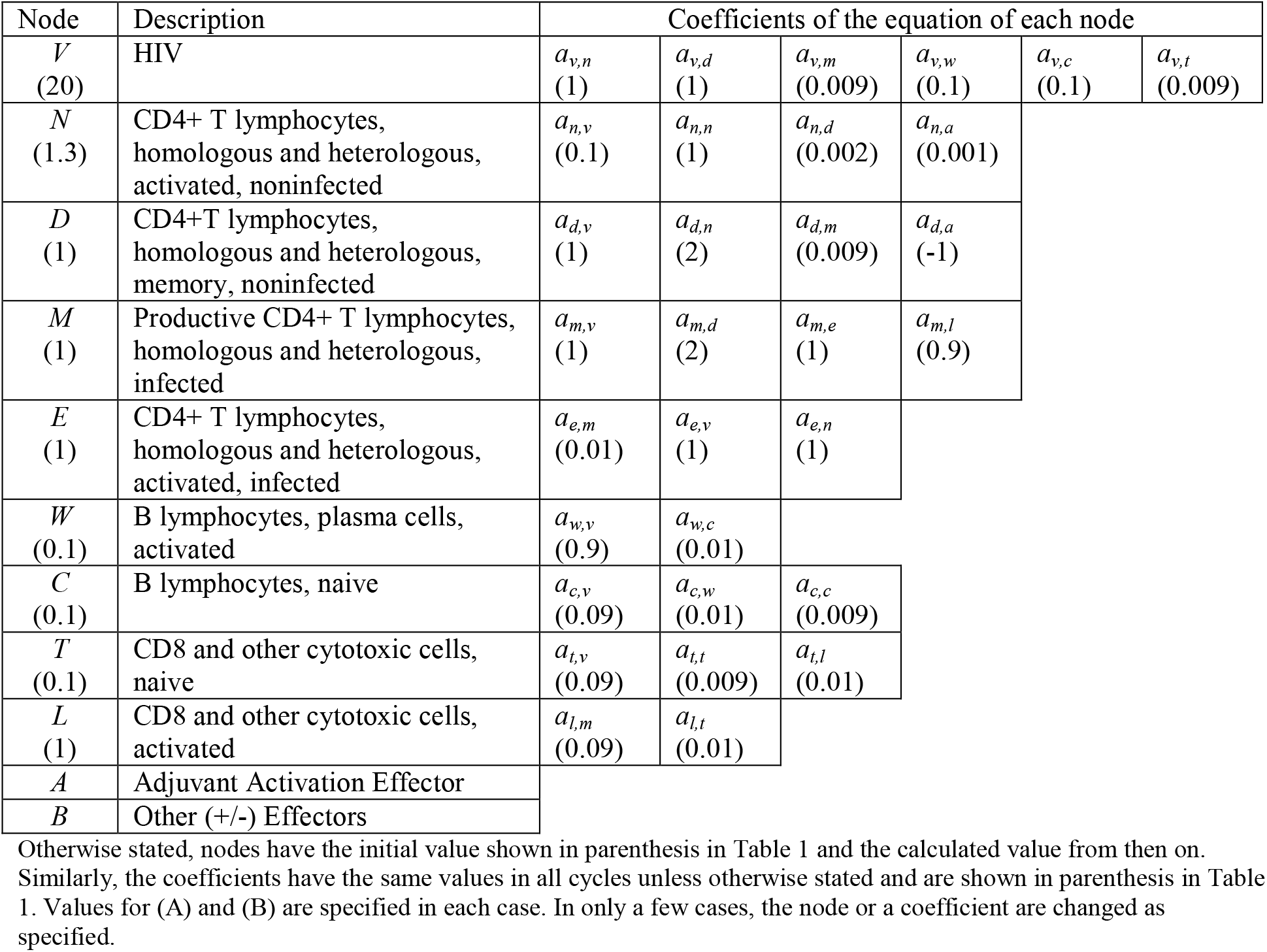
Node description

**Figure 1:**
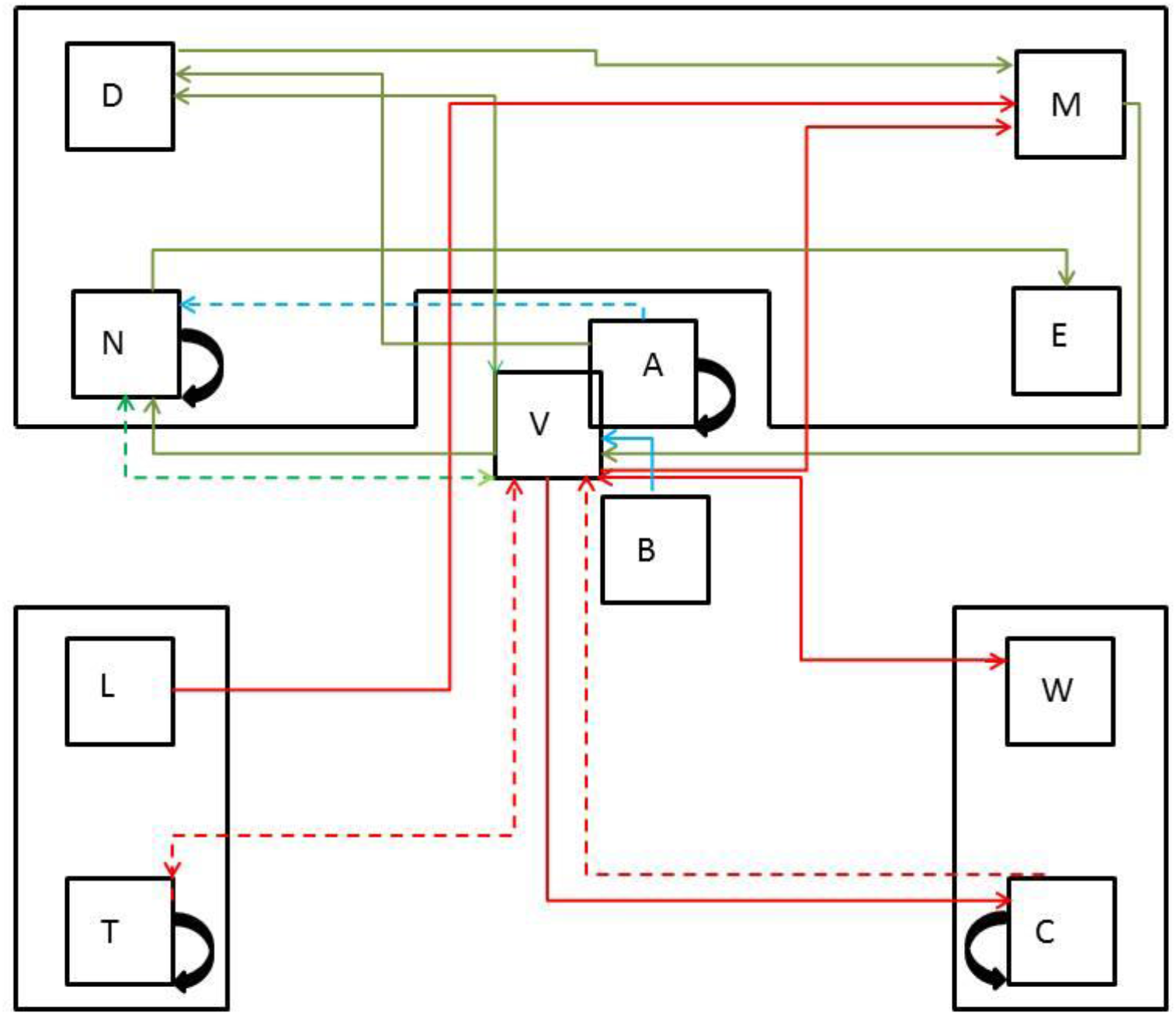
Schematic representation for nodes and their Progressive/Regressive Interaction. Green line, Progressive interactions; Red line, Regressive interactions; Blue line, Interaction with Effectors; Bold line: Direct Interactions; Dashed line, Indirect interactions; Black Curved line, Self interaction. Upper half, CD4+ T cells; Lower half, Left cytotoxic T lymphocites; Right B lymphocites.

### Mathematical approach to the AAE

Using the assumptions from above, we describe the system as 9 coupled differential equations, where each equation gives the rate of change of the virus and of each of the considered cell populations. The node definitions are given in Table 1. These are parameters that constitute the nodes of Figure 1 and the interactions between the nodes correspond with the coefficients of the terms of the differential equations. The coefficients are expressed as *a*_*y,x*_ indicating the activity of y as a function of x. Also, the activated cells have a term for clonal expansion. (A) and (B) are constant (fixed values). The selection of the nodes and the selection of the coefficients implied an optimization of the values, such that only those within a narrow range were acceptable in order to satisfy the differential equations and the designed node system.

Equations

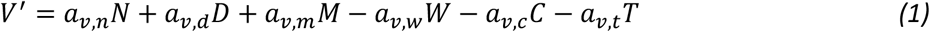

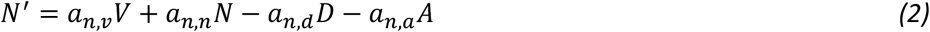

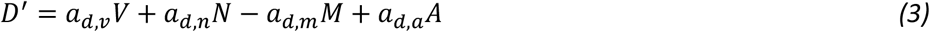

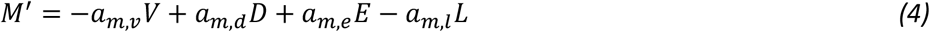

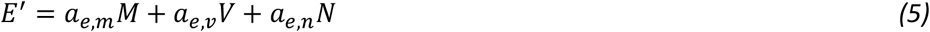

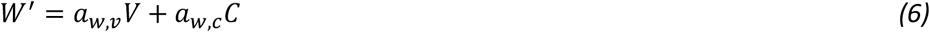

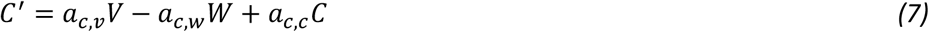

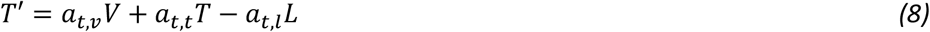

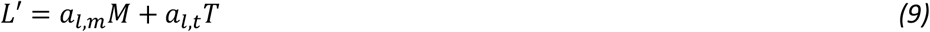

### Excel spreadsheet

The model shown in Figure 1 is the basis for the development of the 9 related differential equations and these are incorporated in the Excel spreadsheet so that after establishing the initial values for the nodes in the first cycle as well as the coefficients for all the equations in all the cycles, then the solution of the differential equations generates the values for the next cycle. This produces a differential progressive (positive) or regressive (negative) value for each node at each cycle in the presence or absence of the AAE, as the system is infected by the virus. The elapsed time is expressed in terms of cycles.

Three considerations were made in order to compare the numbers in the scenarios of infections and recent interventions with those of vaccinations in Africa at the origin of the HIV/AIDS pandemic. First, a consideration of cycles/year was made. In most cases 15 cycles would correspond to a mean of 6 years (a mean of 2.5 cycles/year). This includes the infections and other scenarios discussed in this work although in the case of recent vaccinations such as the STEP trial, the total elapsed time could be less. In the case in which the Origin of HIV infections is discussed in relation to vaccinations in Africa, 1 cycle would correspond to a mean of 2.5 years. Then the ratio of successful interventions of the virus subject to activation in both scenarios (infections and recent interventions in relation to vaccinations in Africa at the origin of the zoonotic infection) is 6.25:1 which indicates that the activations would be 6.25 times more frequently in the present time than at the origin of the virus. This is a maximum value estimated considering only the vaccination events in Africa at the origin of the virus and without taking into account other stimulating activities that could have taken place at that time. Second, with respect to the total elapsed time a mean of 6 years is taken for infections and recent interventions and between 30 and 60 years for vaccination and other activation events in Africa at the Origin of the zoonotic infection. Moreover other factors such as stimulating activities of social, cultural, health, nutritional or of other types, may have occurred for a longer time affecting the time considered as adjuvant activation at the transition from SIV to HIV, giving a ratio of 1:5 (6:30 years) to 1:10 (6:60 years), considering infections and recent interventions to vaccinations in Africa at the origin of the HIV/AIDS pandemic. Third, in terms of the included vaccinated population the ratio would be at least 1:10,000 (e.g. STEP vaccine trial in relation to vaccinations in Africa at the origin of infections) if approximately 3,000 candidates included in the recent vaccination trials up to Clinical Phase II and if more than 30 million individuals in Africa at the origin of the HIV/AIDS pandemic, are considered.

The system is started with an optimized relatively high number for Node V (V=20 Arbitrary Viral Units, AVU) so as to trigger the simulated infection in an Excel spreadsheet with fixed values for nodes and coefficients. The system is designed with a maximum of 21 cycles that simulate the elapsed time. Progressive (positive) and regressive (negative) effects are depicted in Figure 1. This design causes an oscillatory behavior of the values of the nodes between successive cycles, affecting the adjustment of the values to a linear trend, mainly in the absence of the AAE. Moreover, Node V itself was designed so that the virus may cause oscillations in the whole system because it is its own adjuvant effector, even in the absence of other effectors. Also the summation of the values of each node is performed, generating a cumulative curve which adjusts to a linear model. The AAE is programmed as an alternating progressive and regressive effect in order to intervene as discrete non-continuous events. The interaction of the differential equations in the Excel spreadsheet causes a rhythm of events with progressive and regressive effects on the quantitative expression of each node. This simulates the overall progression of the infection over time, the effect of the cellular and humoral control of the infection and most important for the purpose of this work, it simulates the AAE.

### Analysis of the results

The results are sometimes expressed as the value of the node with high adjuvant, (*A*)_*p*_, and with low adjuvant (*A*)_*q*_, as a function of the cycle. The slope (*m*) of the linear regression of each curve is used to define a Progressive/Regressive Index (P/R Index,*I*), as follows:

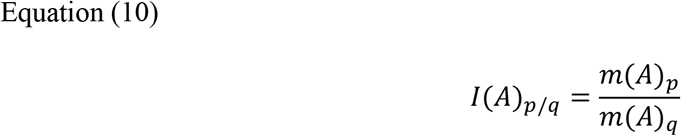

Where *p*>*q*.

The results are also expressed as a cumulative value of the node as a function of the cycle. In this case the summation of the values of the node is calculated as follows:

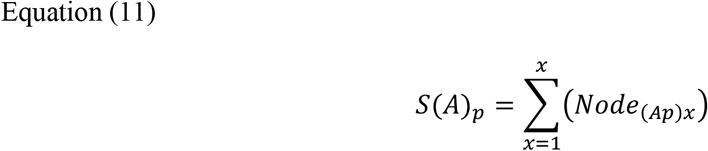

Where (*A*)_*p*_ is the value for the adjuvant and *x* is the number of cycles. Similarly, the relationship or Cumulative Value Index (*SI*) for *S* (A)_*p*_ and *S* (A)_*q*_ is:

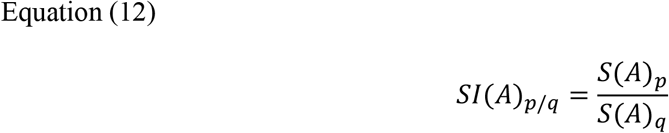

Where *p* > *q*.

In order to quantify the changes in the values of *SI*, they are also expressed as percent, *S* (*A*)_*q*_ / *S* (*A*)_*p*_(100). The obtained value represents percent activation of the condition with the smaller AAE in reference to the condition with the higher AAE. The values of Node V represent Arbitrary Viral Units (AVU) in the organism, whereas the quantification of the other nodes is expressed as Arbitrary Activated Units (AAU).

In some cases, the cumulative value of the node at different values of adjuvant (*A*), is plotted as a function of the cycle showing a linear trend with an *R*^2^which is higher than 0.95. The slope of this curve (*mS*) is plotted as a function of the value of the adjuvant (*A*), in order to show the AAE. The value (*mS*) of is proportional to the potency of node activation. In some cases *mS* is used to calculate the ratio between two conditions (p and q) of (*A*):

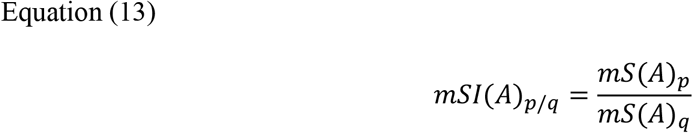

where *p*>*q*.

In order to quantify the changes in the values of *mSI*, they are also expressed as percent,*mS*(*A*)_*q*_ / *mS* (*A*)_*p*_ (100). The obtained value represents percent activation of the condition with the smaller AAE in reference to the condition with the higher AAE and represents the basal level of the potency of activation with low or without the AAE.

The quantitative maximization, the regularity, the symmetry and the refinement of the results of the nodes as a function of time are obtained by choosing values for the nodes and coefficients within a narrow range so that they satisfy the differential equations and their application in the model of Figure 1. Thus the optimization is predetermined by the established design. This leads to two sets of results, one with fixed values for nodes and coefficients and the other with variations in some of the nodes and coefficients as a result of their optimization, as described above.

## RESULTS AND DISCUSSION

### AAE IN THE ABSENCE OF OTHER EFFECTORS (*B*=0)

#### Prediction of the AAE

The AAE hypothesis predicts a differential profile for the viral load as a function of time and for the time required for a collapse of the cellular system, based on the presence or absence of an AAE. It is expected that in the presence of the AAE, the viral load would be greater than in its absence. As shown in Figure 2, both curves, in the absence of adjuvant (*A* = 0) and in its presence (*A* = 3.4) without other effectors (*B* = 0), show an oscillatory behavior which is more pronounced for *A* = 0, after cycle 7, generating a Progressive/Regressive (P/R) profile for Node V, as a function of the cycle. As explained in the Methods section, the *R*^2^of the linear trend may be affected by the P/R oscillations designed in this work, so that the data for *A* = 0 does not fit a linear trend.

**Figure 2.**
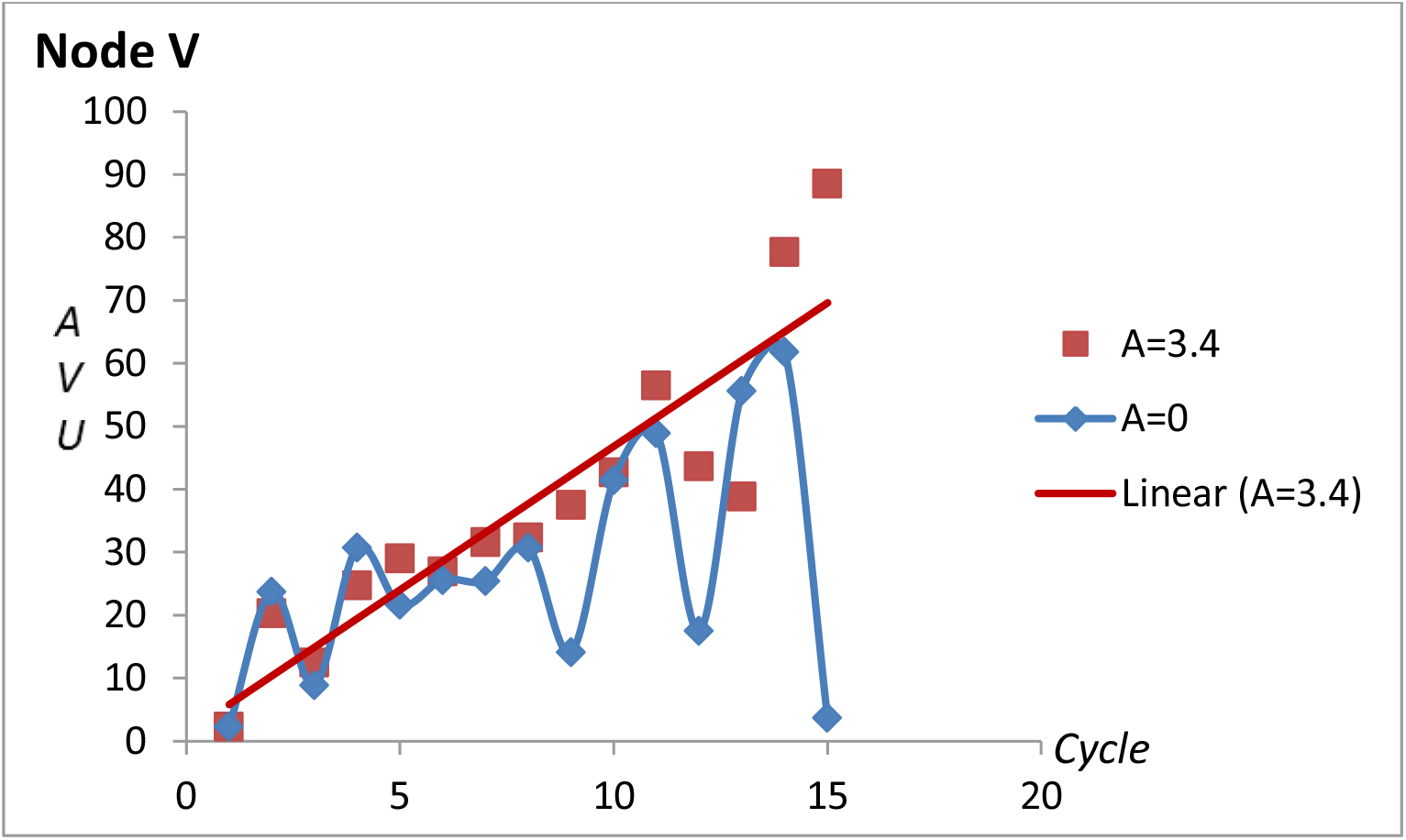
Viral load in each cycle (Node V) up to 15 cycles in the presence (*A* = 3.4) and absence (*A* = 0) of the AAE. The equation for the linear trend for *A* = 3.44 is y = 4.5557x + 1.2494 and *R*^2^ = 0.8052.

The results show a linear trend for Node V in the case that *A* = 3.4 with a slope *m* (*A*)_3.4_ which is 4.56 (*R* ^2^ = 0.81). For *A* = 0 the values are 1.86 and 0.21 respectively, which cannot be used for comparisons due to the low value of its *R*^2^. Therefore, in order to make a comparison of the effect of the AAE, the I(*A*)_*p*/*q*_ was calculated with *A* = 3.4 in reference to *A* = 1 which has a slope *m*(*A*)_*1*_ which is 3.40 (*R*^2^ = 0.63) In this case the P/R Index (Equation 10) is *I*(*A*)_3.4/1_ = 1.34. This shows that the presence of the AAE softens the P/R oscillations on Node V which approaches a linear trend with an increase in the slope and consequently in the viral load.

This effect of the AAE on the P/R oscillations observed on Node V also affects the nodes that represent CD4+ T cells, especially Node M (see below). This shows that the dynamics of the P/R oscillations follow a tight equilibrium in the presence of the AAE and a wide equilibrium in its absence, resembling a stringent control in the former and a relaxed control in the latter. This effect is not observed if the cumulative value (*S*) is considered, showing kinetics which adjust to a linear trend as a function of the elapsed time, whether in the absence or presence of the AAE. This is an interpretation which results from the model designed in this work and does not necessarily represent the Immune System.

Figure 3 shows this effect in a polynomial regression of the same data. Both curves are similar up to cycle 7, after which the presence of the AAE promotes an increase in the values of Node V. In the absence of the AAE, Node V tends to oscillate and then decrease.

**Figure 3.**
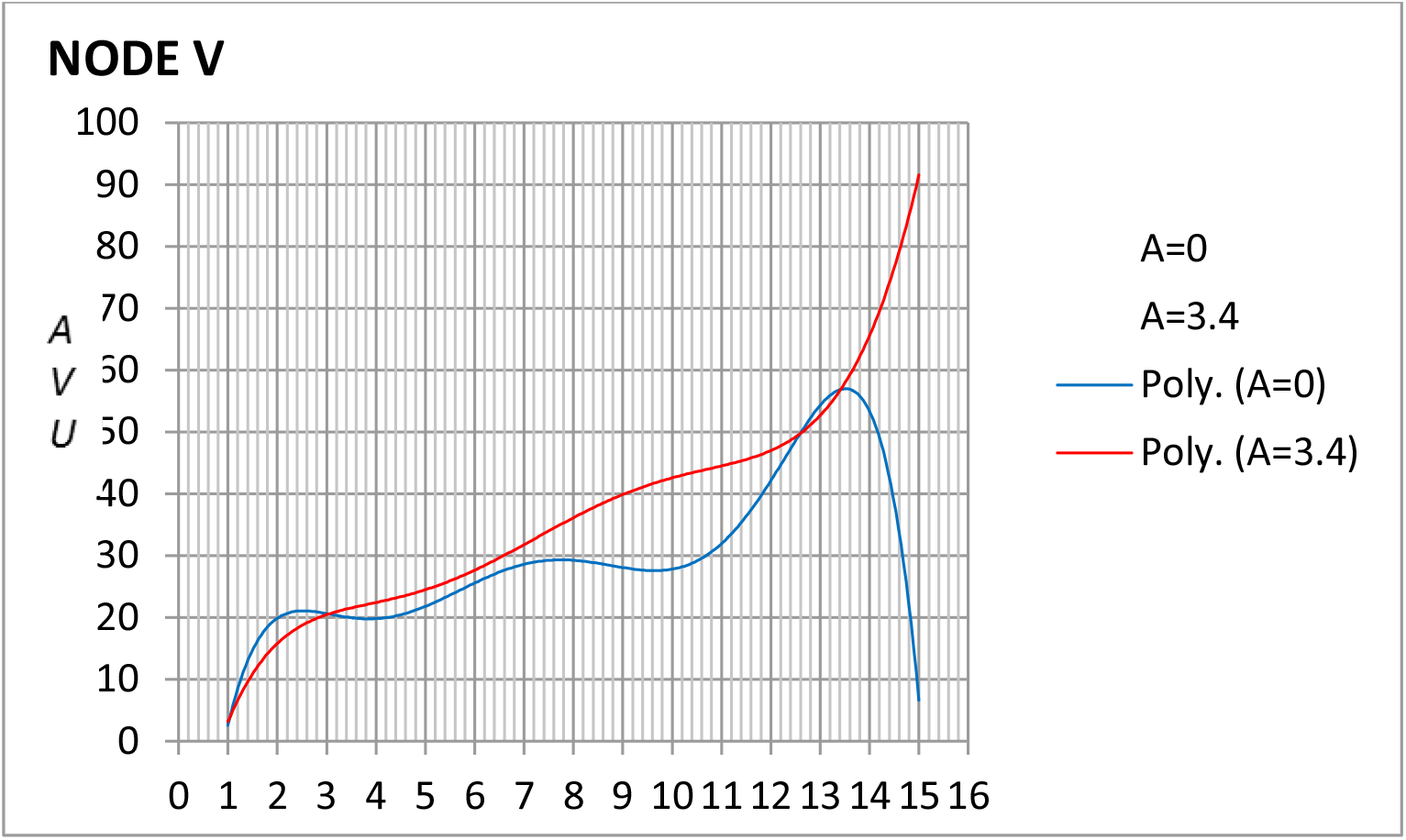
Polynomial regression of the data of Figure 2. The equation for the polynomial trend for *A* = 0 is y = −0.0018x^6^ + 0.0803x^5^ – 1.3921x^4^ + 11.817x^3^ – 51.054x^2^ + 106.27x – 63.182 *R*^2^ = 0.6371 and for *A* = 3.4 is y = −9E-05x^6^ + 0.0077x^5^ – 0.2068x^4^ + 2.4799x^3^ – 14.37x^2^ + 41.283x – 26.094 *R*^2^ = 0.9.127.

Figure 4 shows the cumulative values (*S*) for Node V with A=3.4 and A=0. In contrast to the curves of Figure 2, Figure 4 shows summation curves which adjust to a linear regression each one with an R^2^ which is greater than 0.95 and a slope *m*(*S*) of the linear trend which is 30.18 for *A* = 0 and 38.32 for *A* =3.4.

**Figure 4.**
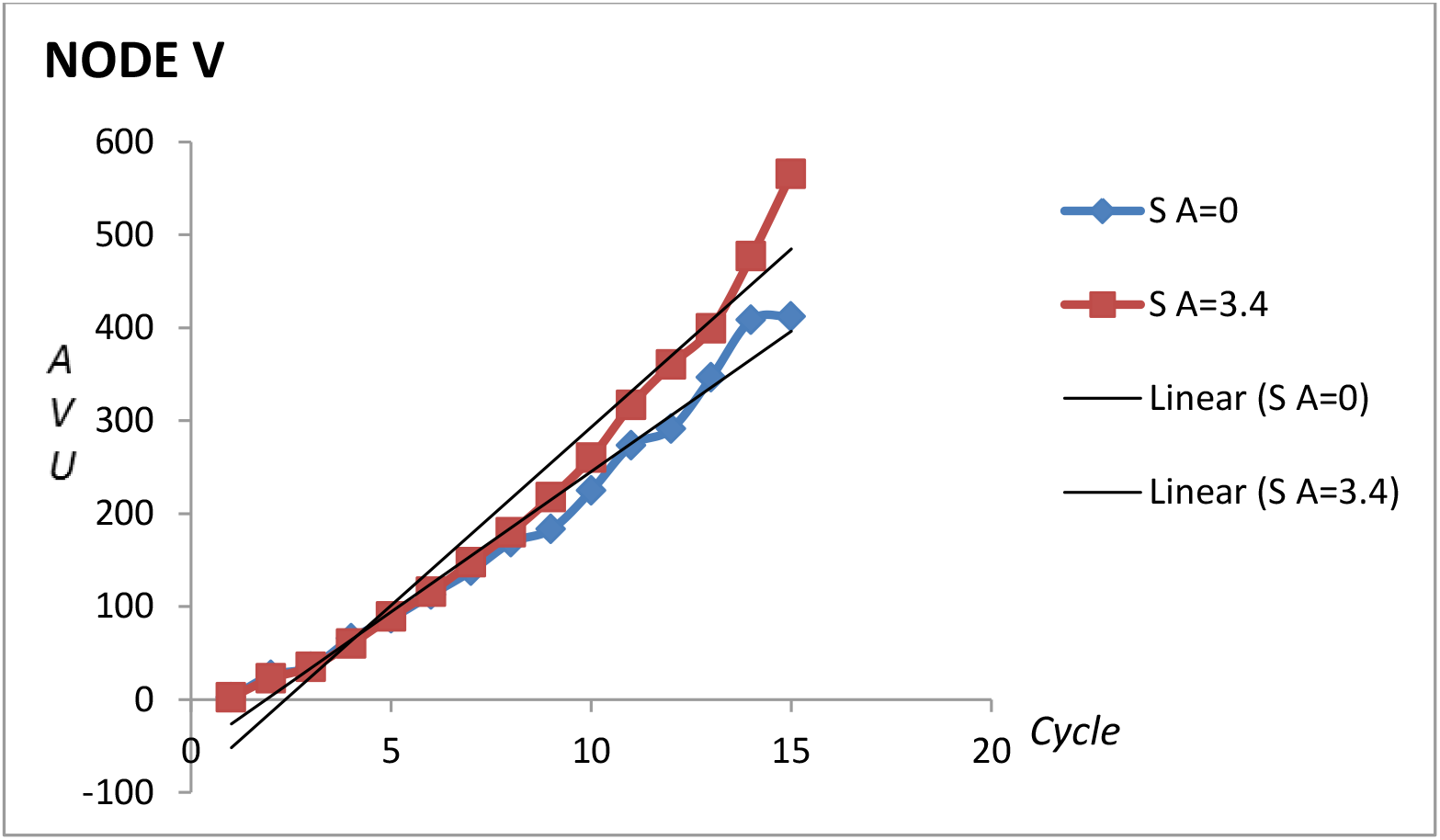
Cumulative value (*S*) of the viral load in each cycle (Node V) up to 15 cycles. The equation for the linear trend for *A* = 0 is y = 30.176x – 56.4 *R*^2^ = 0.9775 and for *A*= 3.4 is y = 38.324x – 90.075 *R*^2^ = 0.9588.

The cumulative value of Node V (*S*) was calculated as the summation of the values of V (Equation 11), in both cases, with and without adjuvant, as follows:

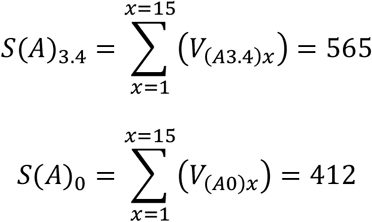

As expected, the AAE causes an increase in the values of Node V, expressed as *SI*(*A*)_3.4/0_ (Equation 12), being 1.37 times higher in the presence than in the absence of the AAE. Considering 15 cycles *S*(*A*)_0_ (412) is 73% of *S*(*A*)_3.4_ (565), indicating that in the absence of the AAE a basal level of virus production is present (see also Table 2).

**TABLE 2.** Cumulative Value Index (*SI*, Equation 12) and Percent activation of each node.

| B=0 | V |  | N |  | D |  | E |  | M |  | C |  | W |  | T |  | L |  |
| --- | --- | --- | --- | --- | --- | --- | --- | --- | --- | --- | --- | --- | --- | --- | --- | --- | --- | --- |
|  | SI | % | SI | % | SI | % | SI | % | SI | % | SI | % | SI | % | SI | % | SI | % |
| $SI(A)_{3.4/0}$ | 1.37 | 73 | 0.99 | 100 | 1.08 | 92 | 1.14 | 88 | 1.76 | 57 | 1.14 | 88 | 1.16 | 86 | 1.18 | 84 | 1.23 | 81 |
| $SI(A)_{3.82/0}$ | 1.33 | 75 | 1.04 | 96 | 1.25 | 80 | 1.15 | 87 | 1.65 | 61 | 1.17 | 86 | 1.18 | 85 | 1.18 | 85 | 1.26 | 80 |

Node V: $SI(A)_{3.4/0} = \frac{565}{412} = 1.37$ and $SI(A)_{3.82/0} = \frac{550}{412} = 1.33$ . %, represents the basal value of percent activation. The values for the other nodes were calculated similarly.

Table 2 shows the result of calculating *SI* and basal percent activation of *A* = 0 for all the nodes of Figure 1. In the absence of any other effector (*B* = 0) and considering a hypothetical case of *A* = 0 for comparison, in the presence of the AAE, with values of *A* = 3.4 (optimized by the slope) and *A* = 3.82 (optimized by *R*^2^), *SI* is nearly 1.0 or slightly higher in most cases and the basal value of percent activation is higher than 50%, indicating that with the assayed values of *A*, the system is activated. This result is in agreement with the hypothesis that the AAE stimulates the nodes. Additionally, this stimulation may favor the infection of Node D by HIV. Node D behaves in a similar way as Node M probably because the virus infects this node to produce Node M. The increase of infections in Node D causes a withdrawal from this node and a decrease in its quantity, increasing Node M which will then decrease by the production of virus and by the cytopathic effect. Then Infection of Node D by the virus converts these cells to Node M, accomplishing the P/R oscillations in both nodes.

The behavior of Node M may be explained by the fact that this is the node that decreases due to the production of virus and to the cytotoxic effect of Node T. Moreover due to the P/R oscillations, this node does not adjust to a linear trend (see Figure 5), as was observed with node V. The value for *SI*(*A*)_3.4/0_ of Node M is 1.76 (765/434, see Figure 6 and Table 2). In this case the basal level of activation of Node M without adjuvant (*A* = 0) is 57% (Table 2).

**Figure 5.**
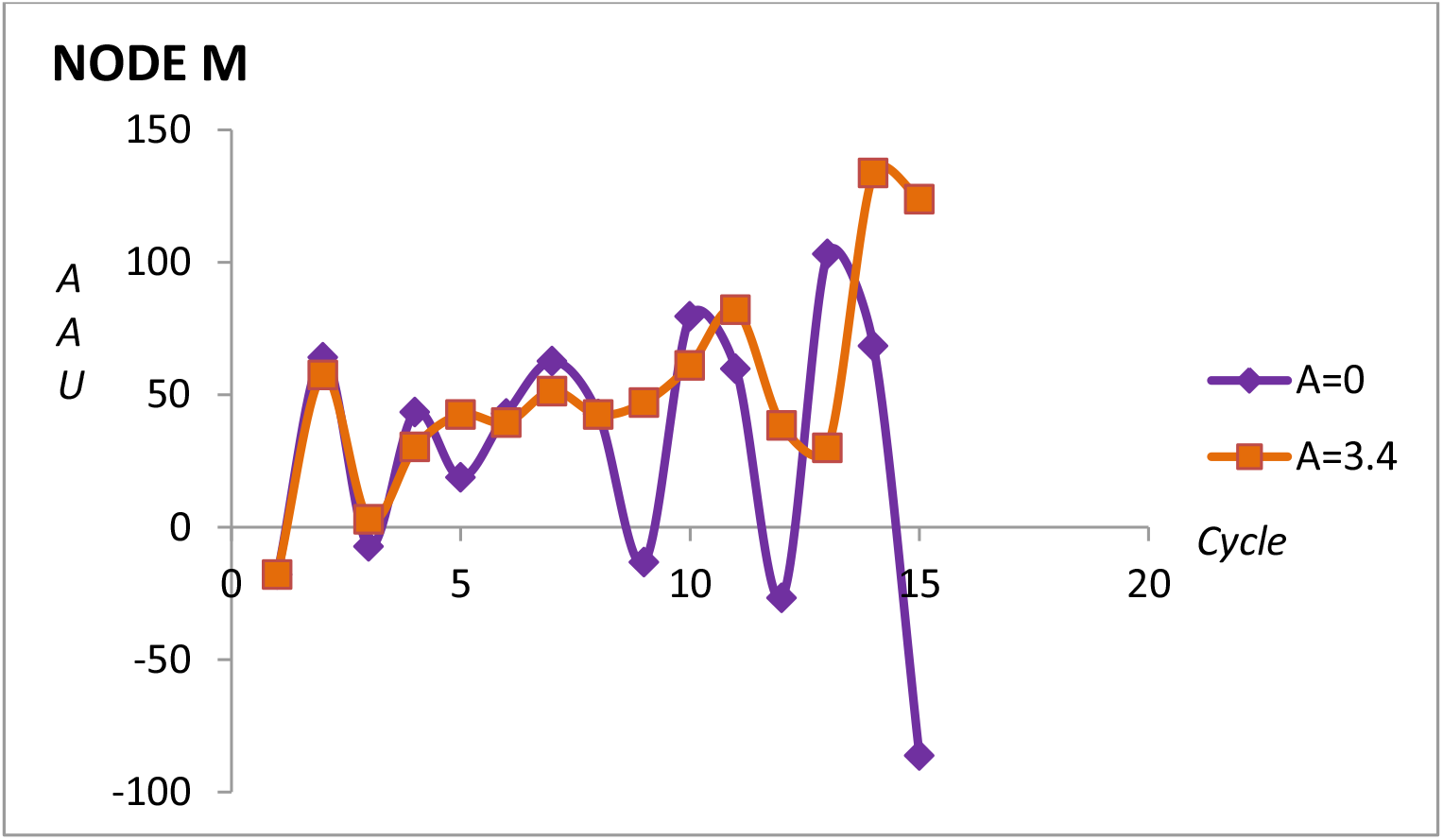
Arbitrary Activated Units (AAU) in each cycle (Node M) up to 15 cycles in the presence (*A* = 3.4) and absence (*A* = 0) of the AAE.

**Figure 6.**
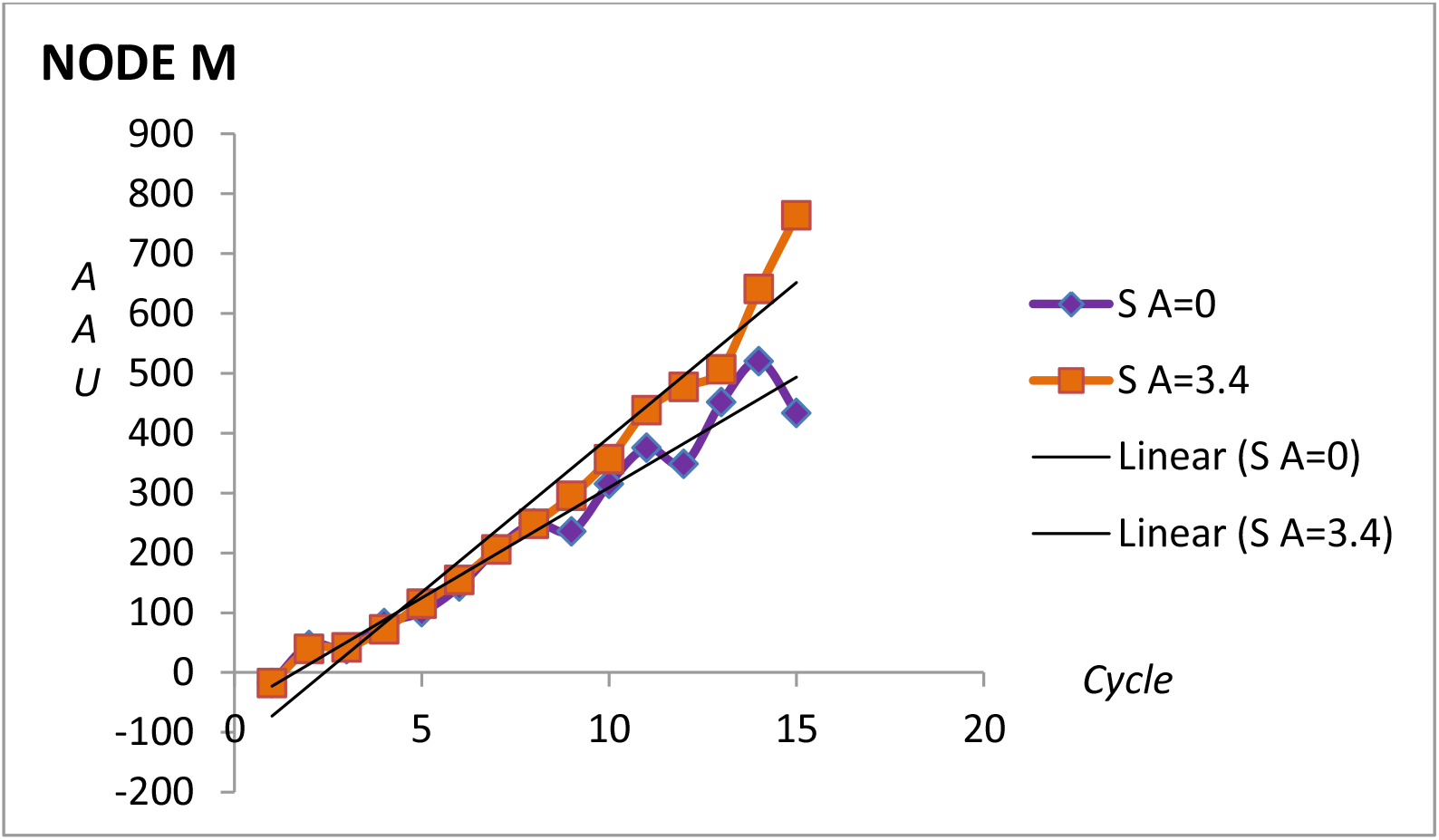
Cumulative value (*S*) in each cycle of Node M up to 15 cycles. The equation for the linear trend for *A* = 0 is y = 36.907x – 59.706 *R*^2^ = 0.9637 and for *A* = 3.4 is y = 51.755x – 124.62 *R*^2^ = 0.9605. *S*(*A*)_0_ = 434, *S*(*A*)_3.4_ = 765, in 15 cycles.

In order to test the AAE on Node V, the cumulative value of the node was plotted as a function of the cycle, showing a linear trend with an *R*^2^ which is higher than 0.95, considering 15 cycles. Then the slope (*mS*) of curves obtained at different adjuvant (*A*) values, were plotted as a function of the value of the adjuvant, in order to show the AAE. Figure 7 shows that *mS* increases as the adjuvant increases within the range 0-3.82, showing the linearity of the AAE. The y-intercept (*A* = 0), basal level for Node V) is relatively high (30.33 AVU/cycle) which is 79% of the *mS* when *A*= 3.82 (38.61 AVU/cycle). This shows the basal level of Node V after the virus stimulates Node N and infects Nodes N and D (see Figure 1) in what is described in the introduction as the dual compromise of the CD4+ T lymphocytes as targets for activation and infection. The presence of the adjuvant increases the slope of the cumulative value of Node V (Figure 7) indicating that as the AAE increases within a certain range (0-3.82), the active production of virus increases up to 1.27 times.

**Figure 7.**
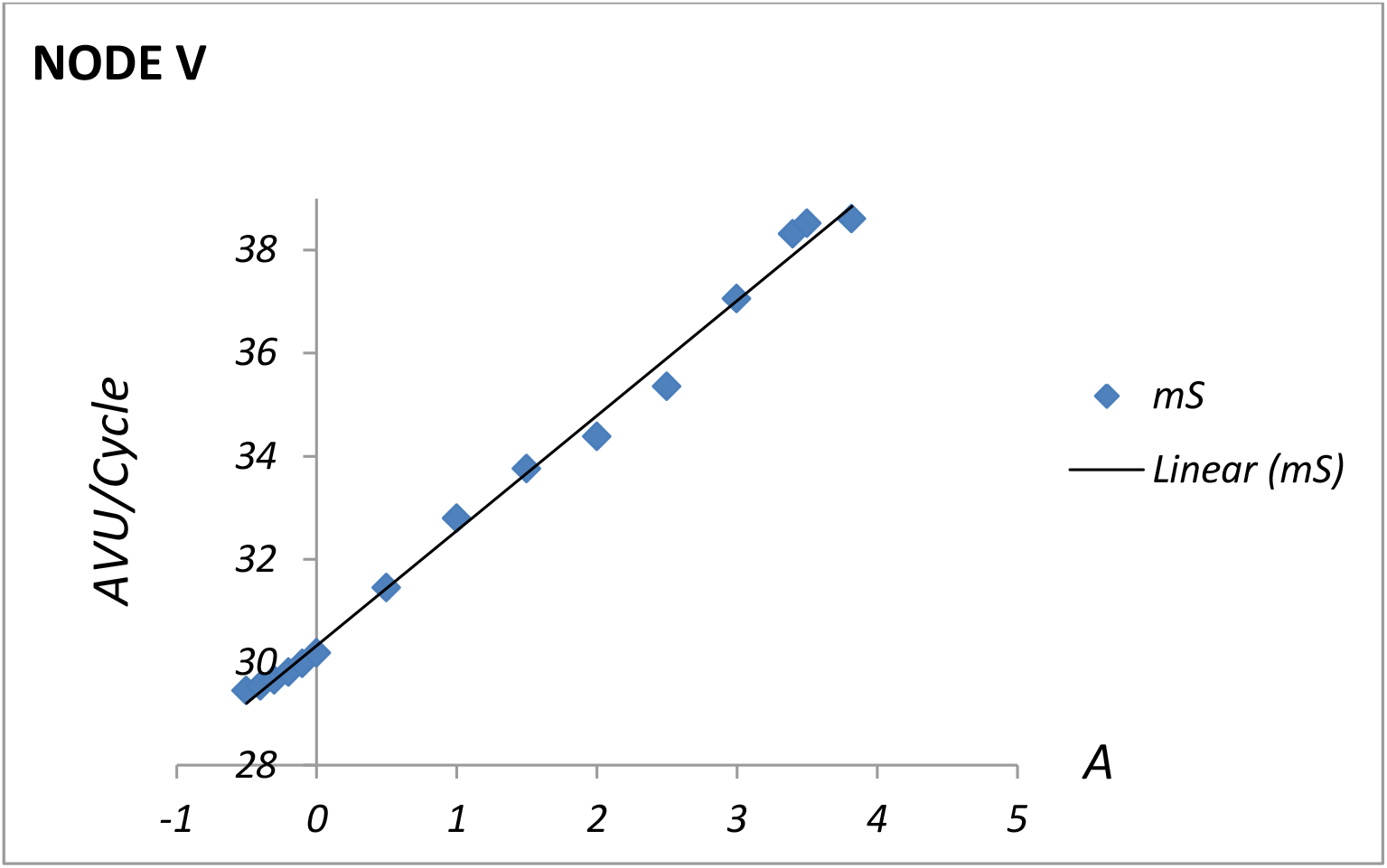
Slope (*mS*) of the plots for the cumulative value of Node V as a function of (*A*) with *B* = 0 in 15 cycles. The last point is *mS* = 38.61 AVU/cycle that corresponds to a value of *A* = 3.82 and to an S(*A*)_3,82_ = 550 AVU. The y-intercept (*mS =* 30.33 AVU/cycle) is the basal level of activation when *A* = 0. The equation for the linear trend is y = 2.2317x + 30.25 *R*^2^ = 0.9943.

#### AAE in vaccine trial failure and in the origin of HIV infections and AIDS development

At this point it is possible to consider these results in the light of the failure of vaccination trials and the origin of HIV infections and AIDS development. In some cases, it has been observed that among the population that enters the vaccination trials, there is a higher tendency for infection in the group of vaccinated individuals than in the group that received placebo. The condition shown as *A* = 0 and *B* = 0 (*A*_0_ *B*_0_) would depict a candidate population which does not suffer an AAE additional to the main activation of the HIV immunogen in the vaccine trial. This means that there will be activation only in response to the viral portion of the vaccine. The condition shown as *A* = 3.82 and *B* = 0 (*A*_3.82_ *B*_0_) would correspond to a candidate population that will have the AAE in addition to the activation due to the HIV portion of the vaccine. This AAE could be caused by different conditions of the population, including the vectors or other components that are incorporated in the vaccine. An example is the STEP (HVTN 502 MRK Ad5) vaccine (O’Connell, Kim, Corey, & Michael, 2012) which was constructed with genes gag/pol/nef on an Ad5 vector. The candidate population that had been previously infected with Ad5, had an immune memory which must have become activated upon receiving the vaccine. The candidate population that received placebo did not activate this immunological memory. When the former population came in contact with HIV, the virus encountered activated cells that were homologous (HIV-specific) and heterologous (e.g. Ad5-specific), but all available for HIV infection. The latter population would only activate homologous HIV-specific cells. It is also possible that other infections and conditions that occur around the timing of vaccination, may promote the AAE.

Figure 8 shows the difference in the potency of activation in terms of *mS*, of Nodes N, D, E and M in 15 cycles with (*A*_3.82_ *B*_0_) and without (*A*_0_ *B*_0_) the AAE (see also Table 3). This could represent the two groups in the example of the STEP trial. Considering that Ad5 infection is widespread within the population and that both groups (vaccinated and placebo) will have memory for Ad5, a higher *mS* is expected to lead to an increased activation of the nodes in the case of the vaccinated individuals (with the AAE) as opposed to the placebo group, with a higher risk of infection in the former than in the latter.

**Table 3.** *mSI* (Equation 13) and Percent activation of each node.

| $B = 0$ | N | | D | | E | | M | | C | | W | | T | | L | |
| --- | --- | --- | --- | --- | --- | --- | --- | --- | --- | --- | --- | --- | --- | --- | --- | --- |
|  | mSI | % | mSI | % | mSI | % | mSI | % | mSI | % | mSI | % | mSI | % | mSI | % |
| Vaccine<br>$A_{3.82/0}$ | 1.06 | 94 | 1.32 | 76 | 1.18 | 85 | 1.39 | 72 | 1.22 | 82 | 1.22 | 82 | 1.22 | 82 | 1.26 | 80 |
| Origin<br>$A_{4.6/0}$ | 2.72 | 37 | 2.23 | 45 | 1.65 | 61 | 2.07 | 48 | 1.62 | 62 | 1.60 | 63 | 1.60 | 63 | 1.60 | 63 |

%, represents the basal value of percent activation.

**Figure 8.**
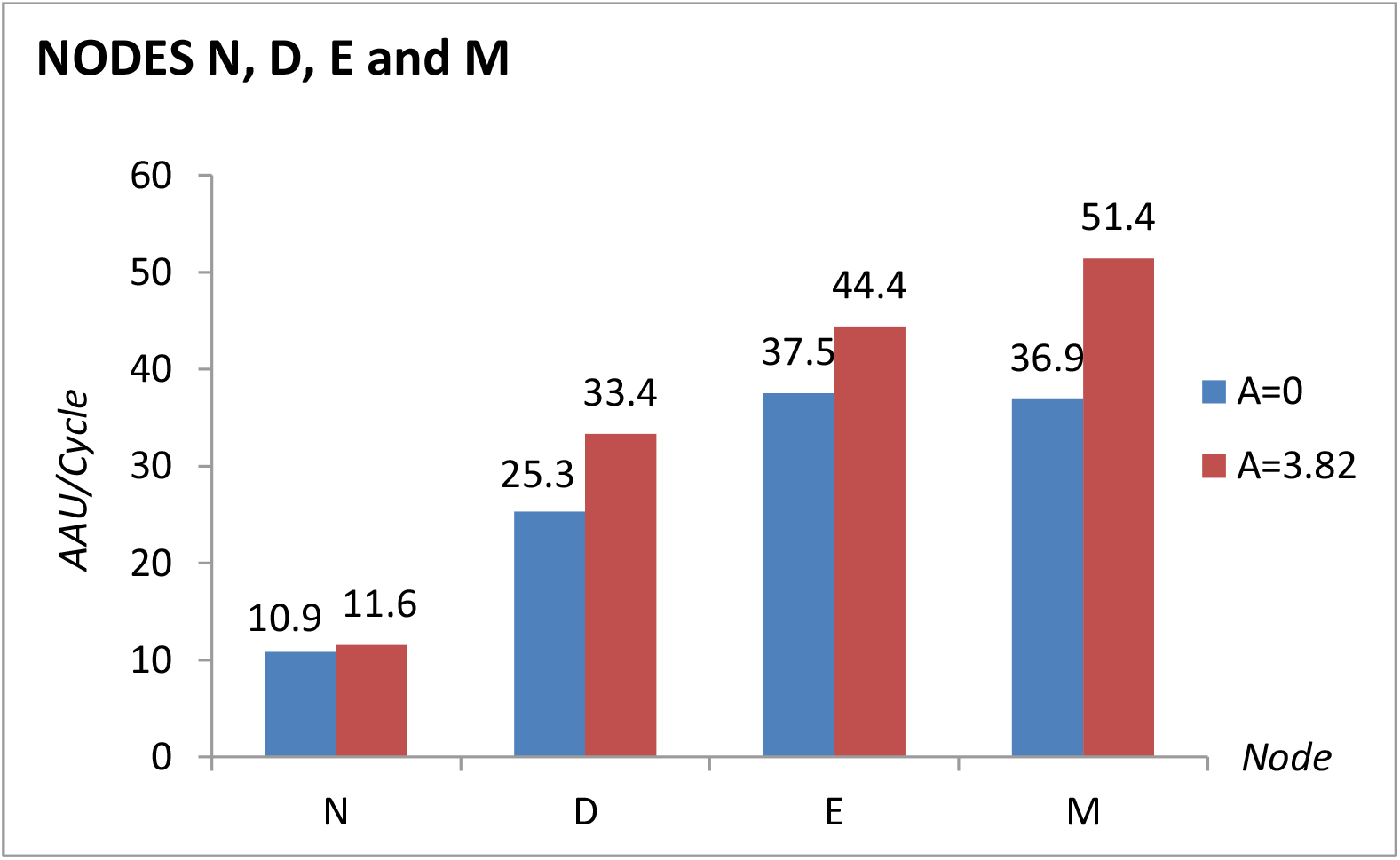
*mS* values in 15 cycles of Nodes N, D, E and M at *A* = 0 and *A* = 3.82.

Figure 9 shows the same response for the nodes that do not represent CD4+ T cells. As expected, the presence of the AAE (*A*_3.82_) increases only slightly the potency for activation of Nodes C, T and L. Node W which represents plasma cells, is higher than the other nodes (see also Table 3).

**Figure 9.**
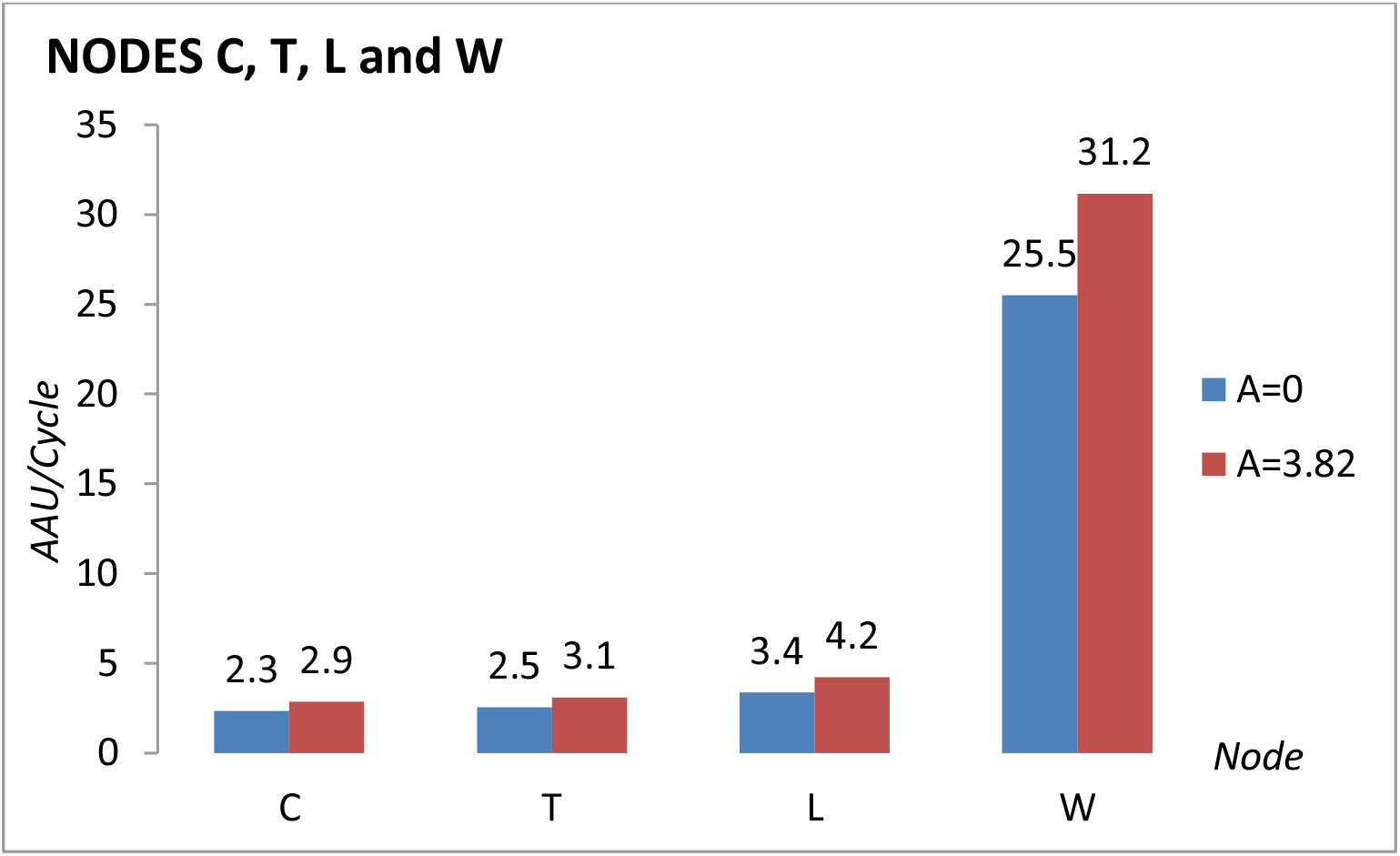
*mS* values in 15 cycles of Nodes C, T, L and W at *A* = 0 and *A* = 3.82.

A similar rationale may be used for the origin of HIV infections and for the origin of AIDS development. In this proposal, in both cases the AAE played an important role in the progression of the infections and of the syndrome. The AAE may have been caused by massive, extensive and intensive vaccinations of more than 30 million people in several countries of Equatorial Africa (Grassi & Andrade, 2001) between the decades 1940-1970, the geographical location where HIV-1 and HIV-2 appeared. At that time, the virus was evolving and adapting to humans and the African human population was challenged by diverse infections and conditions. In order to recreate this scenario, some of the parameters of the differential equations were modified. The initial value of nodes V and N were increased by 35% and 54%, respectively and the coefficient *a*_*n,d*_ of the differential equation for Node N was increased from 0.002 to 0.005. Also the value of (*A*) was set at either 0 or 4.6 and only 12 cycles were considered. Figure 10 shows the difference in the potency of activation in terms of *mS*, of Nodes N, D, E and M in 12 cycles under these conditions chosen for the origin, with (*A*_4.6_*B*_0_) and without (*A*_0_ *B*_0_) the AAE. The results (see also Table 3) show that there is an increase in the potency of activation of Node N (x2.72), Node D (x2.23), Node E (x1.65) and Node M (x2.07) in the former (with the AAE) than in the latter (without the AAE). The effect of at least 6 different vaccines (Grassi & Andrade, 2001) on several tens of millions of people during 30 or more years by activating those cells that were target for the evolving virus may have contributed to its adaptation to humans as a zoonotic infection. Also those vaccines may have triggered the syndrome by having enough opportunities for trial and error for the selection of an adapted HIV and for the emergence of an evolutionary natural positive selective pressure on strains with a high mutation rate. These were adapting to humans, and would have provided a higher number of variant viruses and consequently a higher number of new activatable CD4+ T cells (Grassi & Andrade, 2001).

**Figure 10.**
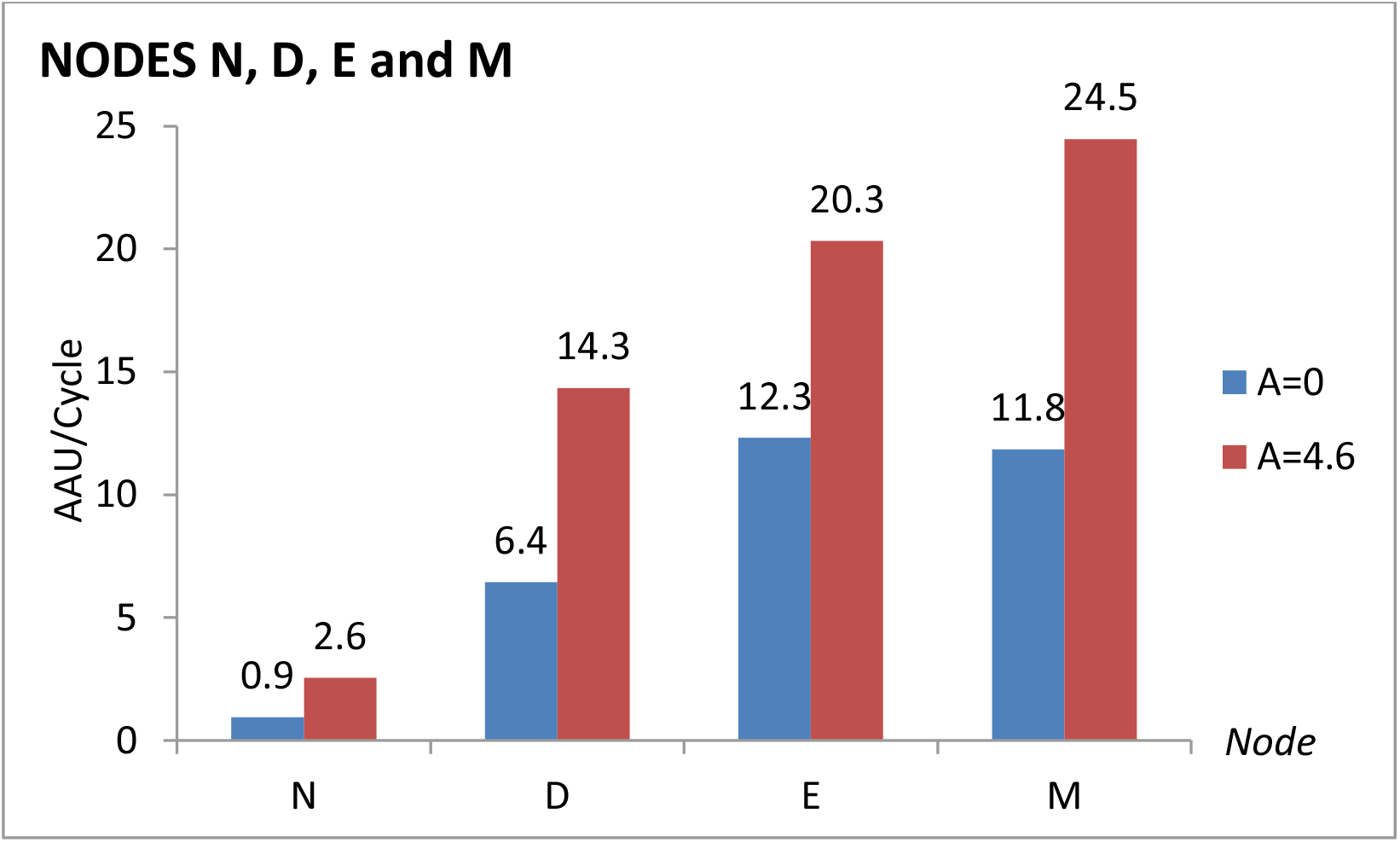
*mS* values in 12 cycles of Nodes N, D, E and M at *A* = 0 and *A* = 4.6.

Figure 11 shows the same response for the nodes that do not represent CD4+ T cells. As expected, the presence of the AAE (*A*_4.6_) increases the potency for activation of Nodes C, T and L. Node W which represents plasma cells, is higher than the other nodes (see also Table 3).

**Figure 11.**
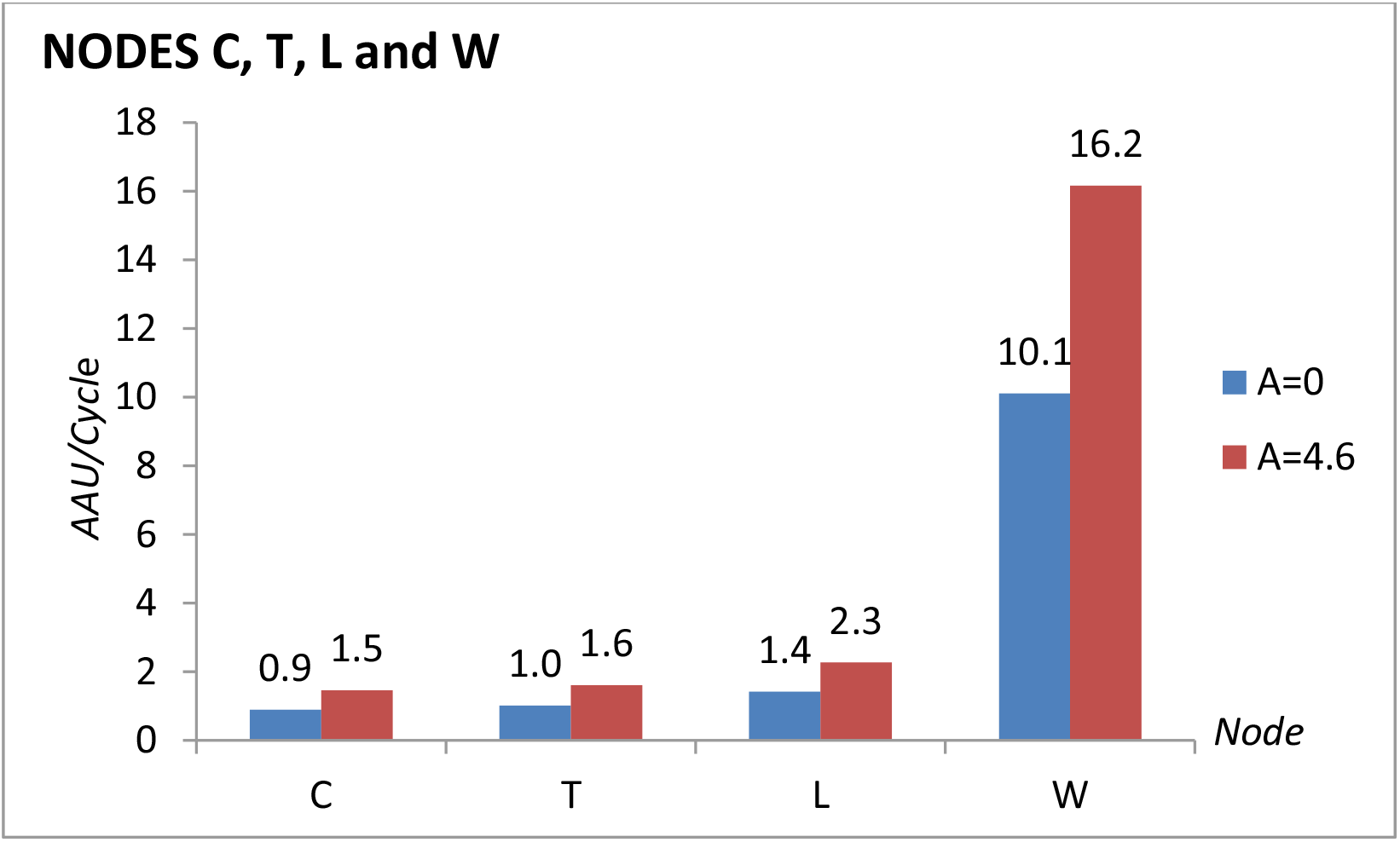
*mS* values in 12 cycles of Nodes C, T, L and W at *A* = 0 and *A* = 4.6.

Considering the cases of vaccine failure and the origins of HIV infections and the AIDS pandemics, two models were designed aiming at the interpretation of each one of these situations with reference to the presence or absence of the AAE. With this in mind the scenarios were set so that in each case, three conditions were met. For the vaccination trials the candidate population was limited in number, it was exposed for a short and limited period of total time and it was organized with many controls and few intervening variables, so that the trial was somewhat in containment. In contrast, for the origin of HIV infections and of the AIDS pandemic, the candidate population involved a higher number of people, for a longer total time and with fewer controls with more intervening variables, it was not in containment and was somewhat chaotic. The populations in these two scenarios are different in their social, cultural, health, nutritional and other characteristics relevant for the AAE proposed in this work. Each one of the scenarios led to a design of our interpretation of events that may represent the conditions with the corresponding results. However, the exact reproduction of the real events is not feasible.

In the model proposed for the origin of HIV infections and of the AIDS pandemic, there is a low basal level for the activation of Nodes N, D, E and M in comparison with the basal level of the vaccination trials that involved a much more controlled population. However, in the presence of the AAE the activation of the same nodes that represent the CD4+ T cells in the exposed African population is much higher than in the vaccination trials. For Nodes C, W, T and L the results are similar but with smaller differences between the basal level and the stimulation by the AAE.

#### AAE in Immunotolerance and Antibody Dependent Enhancement (ADE)

The Adjuvant Activation Hypothesis predicts that adjuvants promote the increase of the value of the nodes that correspond with CD4+ T cells (Nodes N, D, E and M) which in turn promotes the increase of Node V. However, the activation of Nodes N, D, E and M is not always proportional to the AAE. Using this set of differential equations it is possible to try to represent a possible state of Immunotolerance as a function of the AAE. Figure 12 shows the *mS* value of Node V in 15 cycles as a function of adjuvant ranging from 0 to 5.5, with a maximum of *mS* of 38.61 when *A* = 3.82 which then drops to *mS* = 3.5.15when *A* = 5. While the drop in Node V represents an *mS* value of 91% of the maximum value, at *A* = 5 Nodes N, D, E and M reach 100%, 76%, 98% and 81% respectively, if each one is compared with its maximum value of *mS* at *A* = 3.82 This is a comparison of the potency of activation (*mS*) but the actual value at cycle 15 with *A* = 5 of Nodes V and N is 18% and 103% of the value of Nodes V and N at *A* = 3.82 respectively. This resembles the results obtained with Venezuelan patients coinfected with HIV-1 and GB virus C (GBV-C) (Rodríguez, et al, 2014).Then Figure 12 shows that the value of Node V increases as the AAE increases (see also Figure 7) up to a point in which the activation at Node V ceases, approaching the value of Node V when *A* = 0 implying that the system is unable to respond, resembling but not necessarily demonstrating a phenomenon of Immunotolerance. In this case there is a decrease in the response to the adjuvant after a reaching a threshold level of AAE, so that the stimulation of the Cellular Immune System is limited after reaching this threshold affecting the *mS* value of Node V (Figure 12).

**Figure 12.**
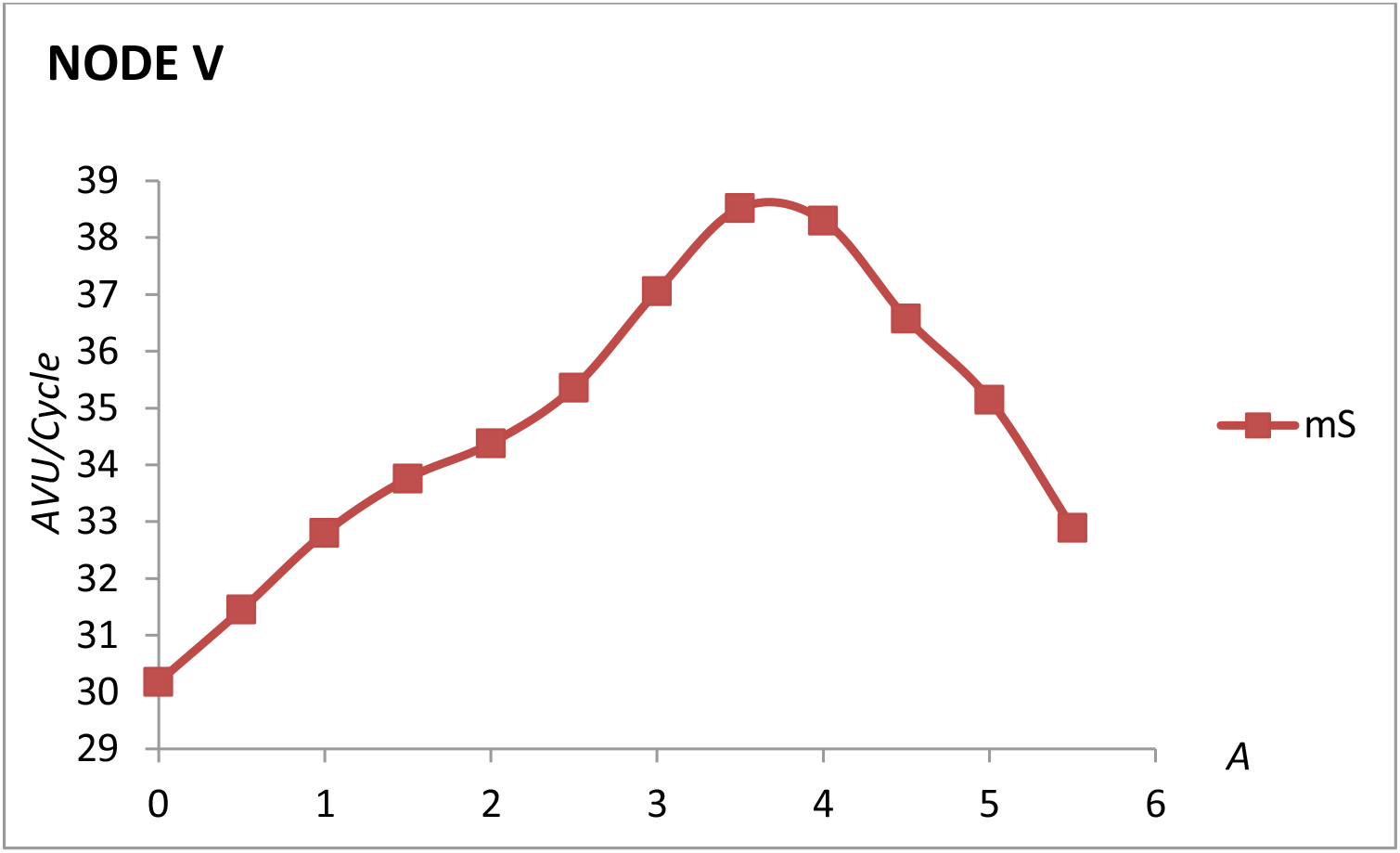
Slope (*mS*) of the plots for the cumulative value of Node V as a function of (*A*) with *B* = 0 in 15 cycles.

In order to create the scenario for a possible Antibody Dependent Enhancement (ADE) (Shmelkov, Nadas, & Cardozo, 2014) of the activity of Node V, the intervention of B cells (Node C and Node W, see Figure 1) was modified so as to increase the effect of this node on Node V. Thus, the term W in the differential equation for Node V (see coefficient *a*_*v,w*_ in Table 1 and differential equation 1) was increased by including a change of sign with an increase in the coefficient of Node W. Then it was possible to evaluate the AAE in the presence of this scenario of ADE and consider the response of Node V to both conditions (AAE and ADE) simultaneously.

Figure 13 shows that upon changing the coefficient *a*_*v,w*_ from 0.1 to −0.3 (thus changing the value of W from negative to positive in differential equation (1)) there is a positive effect on Node V which slightly increases in its values of S *S* and *mS* (maximum value in 15 cycles of the cumulative curve and the slope of the cumulative curve, respectively. See also Table 4. This indicates that an increase in the Humoral Response represented by B cells in terms of Node C and Node W (plasma cells), promotes an increase of Node V (ADE effect) and additionally this response is augmented by the AAE.

**Table 4.** Cumulative (*S*) and *mS* values for Nodes V, C and W as a function of AAE and ADE

| AAE | NODE V |  |  |  | NODE C |  |  |  | NODE W |  |  |  |
| --- | --- | --- | --- | --- | --- | --- | --- | --- | --- | --- | --- | --- |
|  | -ADE |  | +ADE |  | -ADE |  | +ADE |  | -ADE |  | +ADE |  |
| | $S$ | $mS$ | $S$ | $mS$ | $S$ | $mS$ | $S$ | $mS$ | $S$ | $mS$ | $S$ | $mS$ |
| $A = 0$ | 412 | 30.18 | 747 | 51.20 | 36 | 2.34 | 54 | 3.63 | 386 | 25.50 | 590 | 39.60 |
| $A = 3$ | 546 | 37.06 | 915 | 60.39 | 40 | 2.71 | 62 | 4.28 | 435 | 29.56 | 677 | 46.66 |

**Figure 13.**
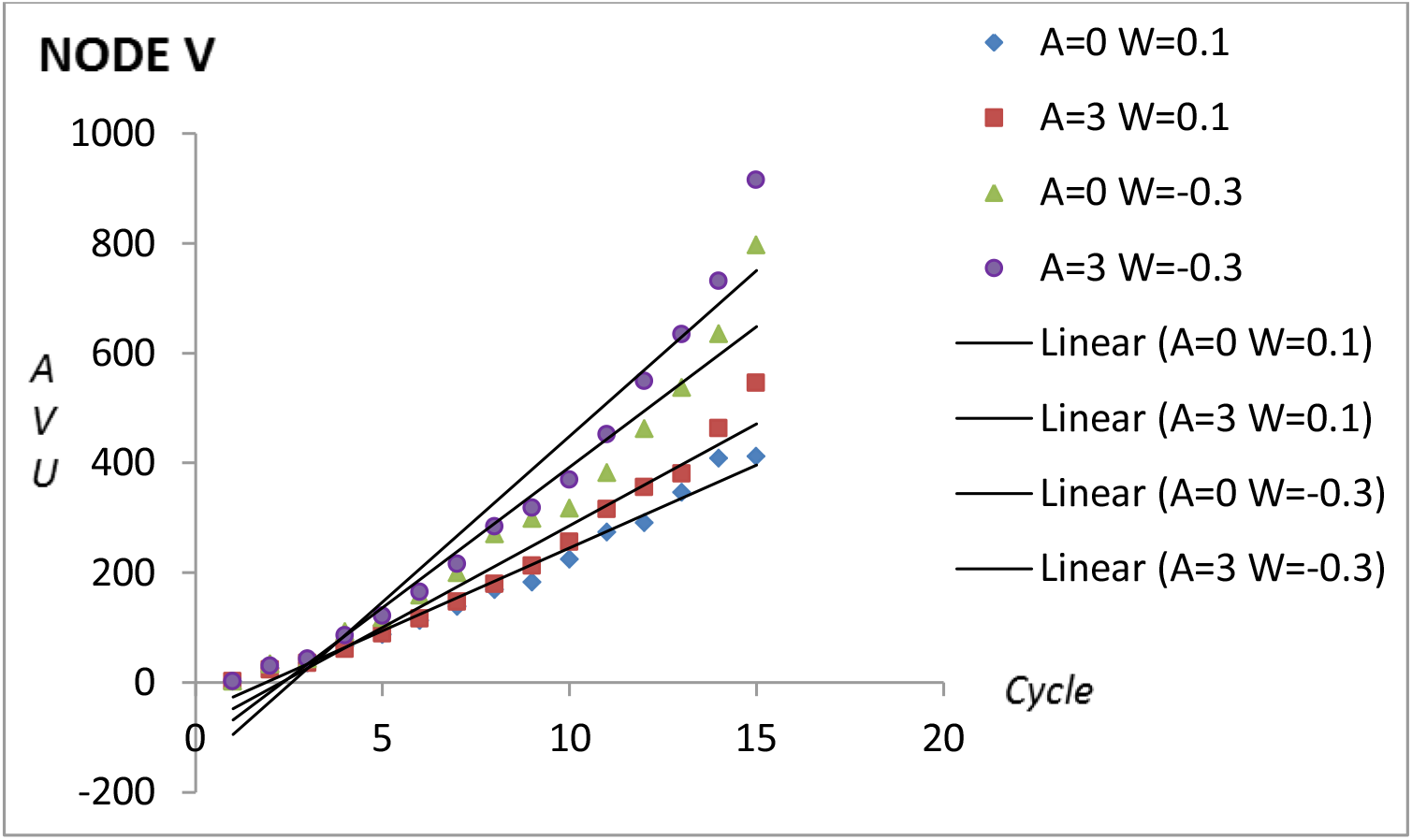
ADE effect on Node V. Cumulative value (*S*) of Node V in 15 cycles with (*A*=3) and without (*A* = 0) adjuvant and with *a*_*v,w*_ =0.1 or *a*_*v,w*_ =-0.3. The equations for the linear trends are y = 60.39x – 155.34 *R*^2^ = 0.9404 for *A* = 3 *a*_*v,w*_ =-0.3; y = 51.203x – 119.71 *R*^2^ = 0.9404 for *A* = 0 *a*_*v,w*_ =-0.3; y = 37.057x – 84.463 *R*^2^ = 0.962 for *A* = 3 *a*_*v,w*_ =0.1; y = 30.176x – 56.4 *R*^2^ = 0.9775 for *A* = 0 *a*_*v,w*_ =0.1.

Table 4 shows two results considering the cumulative value (*S*, maximum value of the cumulative curve) and the potency of the response (*mS*, slope of the cumulative curve) in 15 cycles: First, ADE and AAE on Node V and second, ADE and AAE on the Humoral Response (Node C and Node W). As expected from the result of Figure 13, *S* and *mS* of Node V increase with both conditions, ADE and AAE with a maximum if both effects are present. Node C and Node W behave in a similar way.

#### AAE in the presence of other effectors (B>0)

After describing the value of the basal level of virus production and the value of the AAE, the next step was to try to minimize both values in Node V which with *A* = 3.82 and *B* = 0 add up to *S*(*A*)_3.82_ =550 AVU and *mS* = 38.61 AVU/cycle (see Table 2 and Figure 7). Figure 14 shows the cumulative profiles that reduce each of the effects in 15 cycles. First, with the intervention of an effector (*A* = 6 and *B* = 6.48) the cumulative value of Node V, *S* (*A*)_0_ = 138 AVU and the slope (*mS*) of Node V is 8.91 AVU/cycle which is close to the elimination of the basal level (*mS* = 30.33) AVU/cycle) from the maximum of 38.61 AVU/cycle of Figure 7. Then this first result represents a decrease equivalent to the basal level of virus production (y-intercept of Figure 7). Second, the most obvious way to eliminate the AAE is to consider. This is also shown in Figure 14 (*S*(*A*)_0_ =16.1 AVU when *A* = 0and *B* = 6.48). However, the condition of A=0 is theoretical and is almost unreal for practical purposes. Thus in this work Node V was minimized to almost zero in spite of the presence of the adjuvant, by means of the effectors (*B*). With *A* = 3.1 and B = 7.42 the value of *mS* approaches zero (1E-04 AVU/cycle) and *S* drops from *S*(*A*)_3.82_ = 550 AVU (Table 2) to *S*(*A*)_3.1_ 2.8 AVU (Table 5).

**Table 5.**
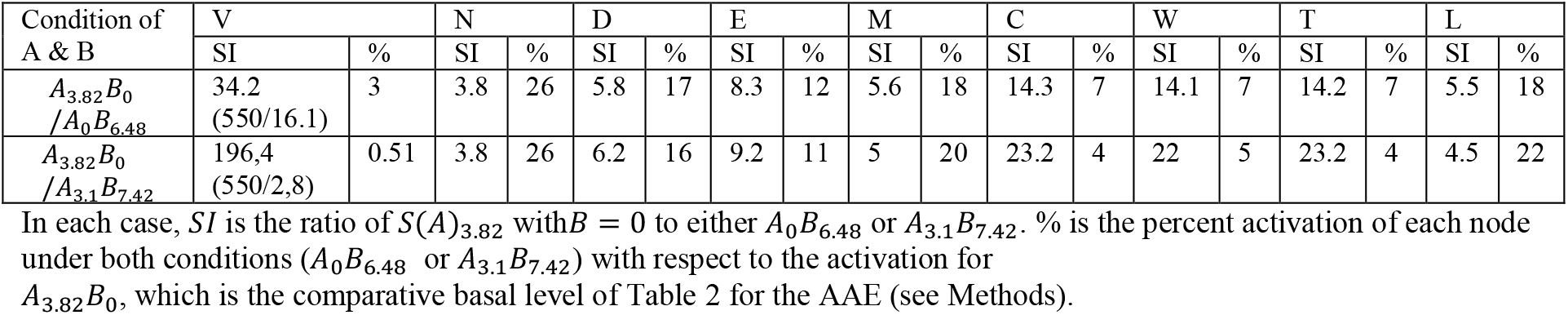
Cumulative Value Index (*SI*, Equation 12) and Percent activation of each node.

**Figure 14.**
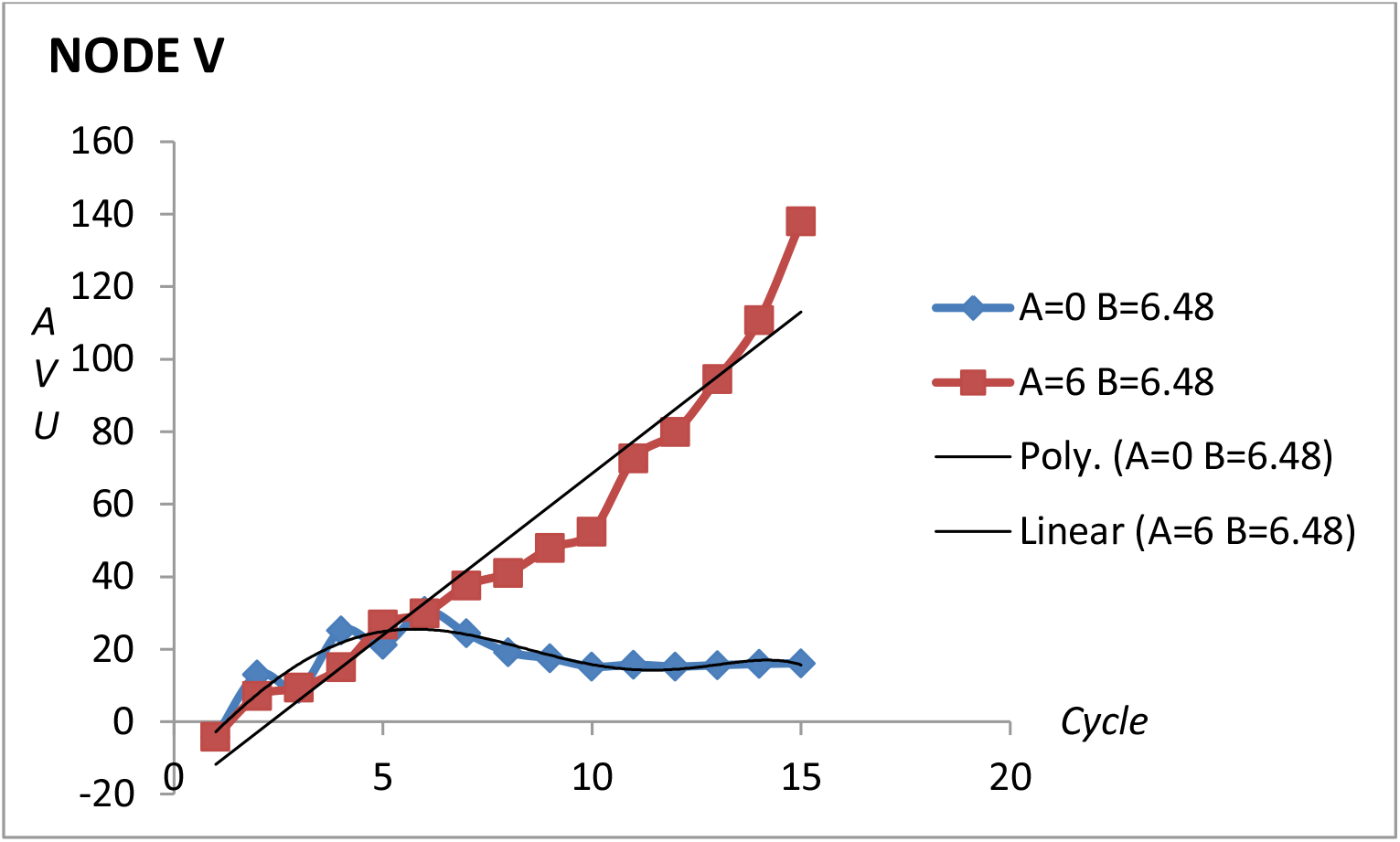
Cumulative value (*S*) for Node V in 15 cycles in the presence (*A* = 6) and absence (*A* = 0) of the adjuvant, and with *B* = 6.48. The equation of the linear trend for *A* = 6 *B* = 6.48 is y = 8.9115x – 20.704 *R*^2^ = 0.94; the equation for the polynomial trend for *A* = 0 *B* = 6.48 is y = −5E-05x^6^ + 0.0011x^5^ + 0.0048x^4^ – 0.1789x^3^ – 0.1677x^2^ + 12.083x – 14.542 *R*^2^ = 0.8378. In 15 cycles for AVU and for *A* = 0 *B* = 6.48 *S* = 16 AVU.

According to Figure 14 the condition *A* = 6 *B* = 6.48 is the best achievement because it shows that it is possible to practically abolish the basal level of of *mS* Node V even in the presence of an adjuvant (*S* = 138 AVU as compared to *S* = 550 AVU for *A* = 3.82) and *B* = 0. Table 5 shows the comparison of the level of node activation between the results of *A*_3.82_ *B*_0_ from Table 2 and the two new conditions introducing an effector B (see Figure 1). The nodes that correspond with CD4+ T cells (Nodes N, D, E and M) decrease in both conditions (*A* = 0 *B* = 6.48 and *A* = 3.1 *B* = 7.42), however Node E is the most affected. This is most likely explained by the fact that the number of infections on Node N decrease due to the decrease of Node V. Then Node E which is the product of infected Node N, decreases and is most affected in its basal level. As expected, the other Nodes that are in close relation with Node V (Nodes C, W and T, see Figure 1) are also affected. With these results it is possible to almost abolish Node V and to minimize the AAE, however this equation system also shows that the numbers and interactions are extremely sensitive to variations. Therefore it is possible to consider the combination of both conditions *A* = 0 from the first row and *B* = 7.42 from the second row of Table 5. This could be suitable for the elimination of Node V and the AAE. Figure 15 shows the plot of the cumulative value for Node V under these conditions (*A* = 0 *B* = 7.42), indicating that in 8 cycles the tendency approaches zero with a maximum of approximately 20 AVU in cycle 4. Under these conditions and if the plot includes 15 cycles, the tendency is strongly negative (*S*(*A*)_0_ = − 980 *AVU* and *mS* = −38.29 AVU/cycle). This is a condition which may be very difficult to interpret and also almost impossible to put in practice. However, it could be possible to manage both effects by controlling simultaneously immune activation, *A* = 0 simulating immune therapy, immune suppression or passive immunization (Grassi & Andrade, 2001) and minimizing Node V with anti-retroviral therapy, *B* = 7.42 (Goldstein & Chicca, 2012).

**Figure 15.**
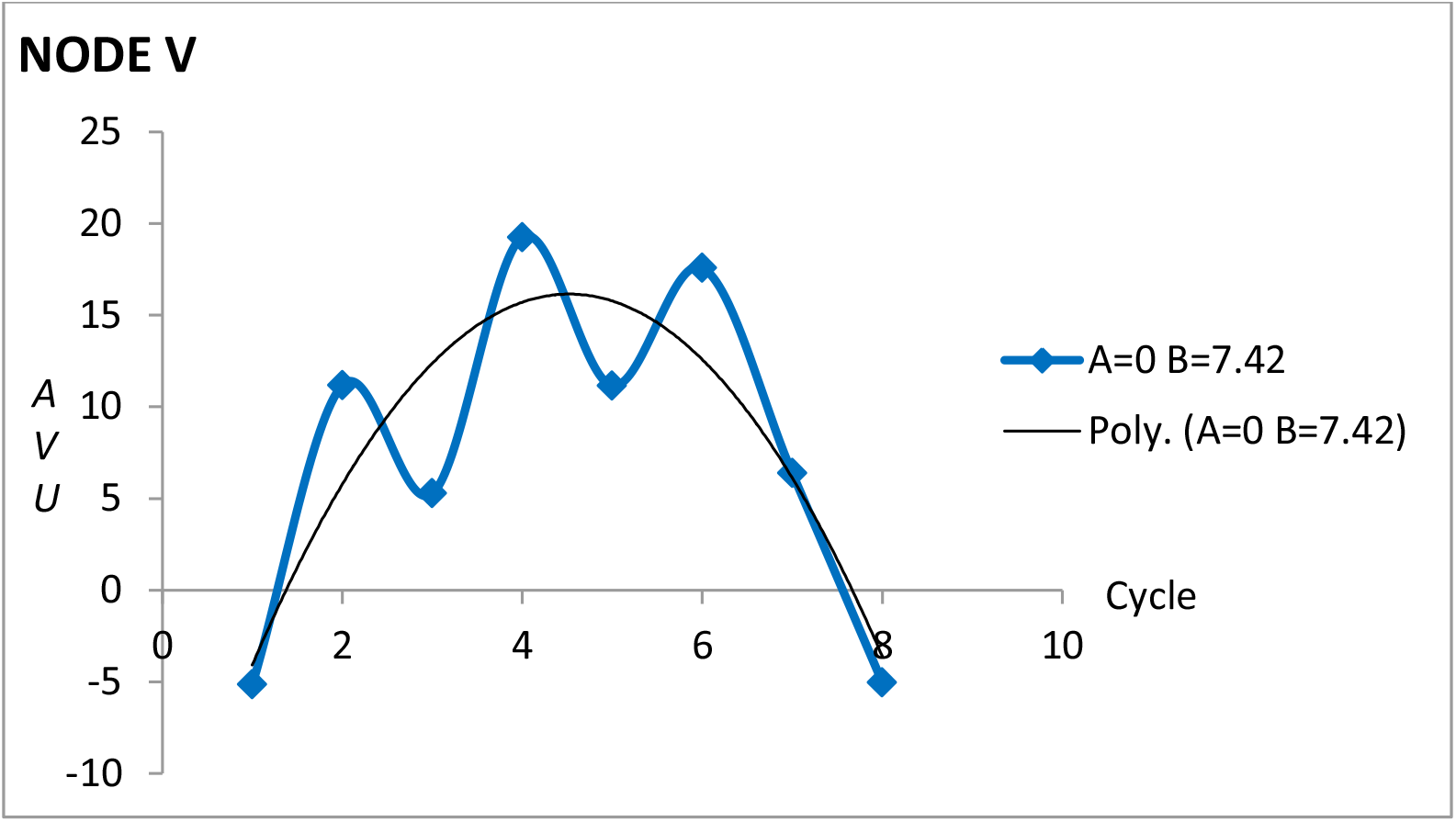
Cumulative value (*S*) for Node V in 8 cycles with = 0 *B* = 7.42. The equation for the polynomial trend is y = −1.6316x^2^ + 14.75x – 17.19 *R*^2^ = 0.7595.

In these cases production at Node V may be affected with either one of two conditions: with *A* = 0 or with relatively high values of an effector *B*. The absence of *A* means that there are no stimulations of the Immune System. Then the former (*A* = 0) could be considered as a theoretical condition because there is a very low probability for an organism to maintain a low-stimulation state under real conditions. Otherwise, this could be achieved by some condition of immune suppression. The latter implies that the involvement of CD4+ T cells in the dual compromise as helper-activatable cells/targets of viral infection has to be impaired. According to the results it may be interpreted that the basal levels reported for Node M (Figure 6) and for Node V (*mS* in Figure 7) are contributing to this dual compromise of activation/infection. Therefore, the therapeutic target could be to affect the basal level of these two nodes. The impairment of reproduction of the virus has already been achieved with anti-retroviral therapy (ART). The challenge is to impair Node M which is the product of the activation of Node N, the infection of Nodes D and E and the transition to a memory state. It would be desirable to have viruses or their parts attenuated for the activation function, but these have not been described. Then, since organisms have many ways to suffer the AAE, the desired state is one which combines the absence of adjuvants and the presence of effectors that control virus reproduction (such as ART) or eliminate it (such as passive immunization or broad mAb therapy). What is clear is that it is necessary to decrease the dual function of CD4+ T cells. This includes decreasing the virus but also, decreasing the AAE.

## CONCLUSION

This work does not present an exhaustive analysis of the human Immune System nor does it criticize the use of vaccines as a design for the prevention of infections. Instead it analyzes the hypothesis of the AAE and its consequences at the origin of the HIV/AIDS pandemic and in the failure of HIV vaccine trials. The model of cell interactions, the differential equations and the coupling of those differential equations to the Excel spreadsheet have led to the analysis of the AAE as well as of its hypothetical participation at the origin of the virus and at the present day vaccine trials and failures. Other effects that resemble immunotolerance and ADE, as well as the use of passive immunization and anti-retroviral therapy, were evaluated.

With this proposal it was possible to increase the value of Node V (viral load) in more than 30% and the potency of activation in more than 25%, in response to the presence of the adjuvant in an evaluation of the AAE. Furthermore, the intervention of the AAE at the origin of the HIV/AIDS pandemic was shown in this model as well as its correlation with the failure of vaccine trials.

For future studies the simulation presented in this work may be adapted to evaluate other variables such as the thermodynamic behavior of the HIV infection and the characterization of its different stages (González, Figueiredo, & Coutinho, 2020).

## Data Availability

All data is available

## ACKNOWLEDGEMENTS

The authors acknowledge Professor Magdiel Ablan, Ingeniería de Sistemas, Facultad de Ingeniería, ULA, for the request of advice for her students Yonny Fernández, Carla Rangel and Orielis Osorio in the context of the course Simulation, allowing the first considerations on this matter. This work was financed by the authors.

## AUTHOR CONTRIBUTION

HCG, EDJA, conceived and designed the analysis and the interaction of the nodes; EDJA, developed the differential equations; HCG designed the Excel spreadsheet and wrote the paper; VJA-G, performed calculations to obtain the quantitative maximization, the regularity, the symmetry and the refinement of the results; EDJA, VJA-G, performed the revision and correction of the paper.

